# Imaging Sensitive and Drug-Resistant Bacterial Infection with [^11^C]-TMP: *In Vitro* and First-in-Human Evaluation

**DOI:** 10.1101/2021.09.21.21262899

**Authors:** Iris K. Lee, Daniel A. Jacome, Joshua K. Cho, Vincent Tu, Anthony Young, Tiffany Dominguez, Justin D. Northrup, Jean M. Etersque, Hsiaoju Lee, Andrew Ruff, Ouniol Aklilu, Kyle Bittinger, Laurel Glaser, Daniel Dorgan, Denis Hadjiliadis, Rahul M. Kohli, Robert H. Mach, David A. Mankoff, Robert Doot, Mark A. Sellmyer

## Abstract

Recently, several molecular imaging strategies have developed to image bacterial infections in humans. Nuclear approaches, specifically positron emission tomography (PET), affords sensitive detection and the ability to non-invasively locate infections deep within the body. Two key radiotracer classes have arisen: metabolic approaches targeting bacterial specific biochemical transformations, and antibiotic-based approaches that have inherent selectivity for bacteria over mammalian cells. A critical question for clinical application of antibiotic radiotracers is whether resistance to the template antibiotic abrogates specific uptake, thus diminishing the predictive value of the diagnostic test. We recently developed small-molecule PET radiotracers based on the antibiotic trimethoprim (TMP), including [^11^C]-TMP, and have shown their selectivity for imaging bacteria in preclinical models. Here, we measure the *in vitro* uptake of [^11^C]-TMP in pathogenic susceptible and drug-resistant bacterial strains. Both resistant and susceptible bacteria showed similar *in vitro* uptake, which led us to perform whole genome sequencing of these isolates to identify the mechanisms of TMP resistance that permit retained radiotracer binding. By interrogating these isolate genomes and a broad panel of previously sequenced strains, we reveal mechanisms where uptake or binding of TMP radiotracers can potentially be maintained despite the annotation of genes conferring antimicrobial resistance. Finally, we present several examples of patients with both TMP-sensitive and drug-resistant infections in our first-in-human experience with [^11^C]-TMP. This work underscores the ability of an antibiotic radiotracer to image bacterial infection in patients, which may allow insights into human bacterial pathogenesis, infection diagnosis, and antimicrobial response monitoring.

**One Sentence Summary:** The PET radiotracer [^11^C]-trimethoprim shows high uptake in both TMP-sensitive and -resistant bacteria *in vitro*, the potential for imaging many different pathogenic strains, and uptake in patients with active infections.

## Introduction

The ability to specifically detect and characterize a bacterial infection in a patient has been a long-sought goal for molecular imaging (*1*). The implications of such a technique are wide-ranging and could improve diagnosis of bacterial infection as well as allow quantitative monitoring of the effects of antimicrobial treatment. The current standards of biopsy and *ex vivo* microbial culture are hampered by contaminations, sampling limitations, variable sensitivity, potential for procedural complications, and delayed results (*2*). Thus, bacterial specific radiotracers could have positive impact on clinical practice and move the field of infection imaging beyond the current practices which generally apply non-specific nuclear imaging approaches such as radionuclide-tagged white blood cell (WBC), Gallium-67 citrate, and [^18^F]-fluorodeoxy-glucose (FDG) scans (*3, 4*).

To develop bacterial specific radiotracers, several general approaches have been pursued. One is targeting biochemical/metabolic transformations that are unique to bacterial biology. For example, several groups have made advancements in preclinical models using radiotracers including [^18^F]-fluorodeoxy-sorbitol (FDS), [^11^C]-para-aminobenzoic acid (PABA), D-amino acids, and probes targeting the maltose transporter in bacteria (*5-8*). While these results in animals have been promising, only [^18^F]-FDS has been reported in human patients at this time (*9*). Another strategy is to use radiolabeled antibiotics which have inherent selectivity for bacteria over human cells. We previously reported the development of [^11^C]- and [^18^F]-labeled trimethoprim (TMP)-based radiotracers and have demonstrated uptake specificity to bacterial infection over other pathologies such as sterile inflammation (turpentine) and neoplasm (breast carcinoma) in rodent models (*10, 11*). As opposed to metabolic radiotracers, those radiotracers that build on antibiotics such as TMP have raised concerns that imaging antibiotic resistant bacteria may be challenging. Furthermore, bacterial infection radiotracers need to maintain high uptake across a broad spectrum of bacterial species (e.g. targeting both gram-positive and gram-negative species) to produce favorable positive and negative predictive test characteristics when in the causative organism is as yet unknown (*12*).

Here, using previously tested laboratory bacterial strains and new, comparable, drug-resistant clinical isolates, we performed dose-response assays with a variety of antibiotics with specific attention to TMP resistance. We then tested uptake of [^11^C]-TMP in these pathogenic species to determine whether TMP resistance by itself is a critical factor for the “imageability” of the bacterial species. In addition, we performed whole genome sequencing (WGS) of the clinical isolates to identify the number of dihydrofolate reductase (DHFR) genes and to probe for other resistance mechanisms present in these strains. To extend our WGS results, we used the NCBI database of annotated whole bacterial genomes and assessed the top 19 pathogenic bacterial species for the presence of wild-type and mutant DHFR enzymes. With evidence of the ability to image both susceptible and resistant strains, we report the first-in-human study of bacterial infections that are imaged with [^11^C]-TMP, with special attention to lung and spine infections. We discuss cases that demonstrate the biodistribution and specificity of [^11^C]-TMP uptake, and include instances when antibiotic treatments were administered to highlight the potential of [^11^C]-TMP to add valuable diagnostic information in different clinical cases of bacterial infection.

## Results

[^11^C]-TMP was synthesized as previously reported (**Fig. 1A**, (*11*)). We previously tested TMP-sensitive bacterial strains in animal models (*10*), and thus TMP-resistant clinical isolates of the same species were acquired (details included in the **Materials and Methods** section). Resistant strains from the clinical lab were known to be resistant to TMP/SMX based on CLSI M100 breakpoints. To validate the susceptibility or resistance of our bacterial panel to TMP alone, we performed dose-response assays. Each bacterial strain was incubated with varying concentrations of TMP (0-50 μM) for 6 hours (**Fig. 1B**), growth endpoints were recorded, and IC_50_ values were calculated (**Fig. 1C**). We classified each strain as either susceptible or resistant to TMP based on both the qualitative observation of the dose-response curve and the quantitative IC_50_ value obtained. All bacterial strains were further profiled for resistance against other antibiotic classes to characterize multi-drug resistance. (**Fig. S1**).

**Figure 1.**
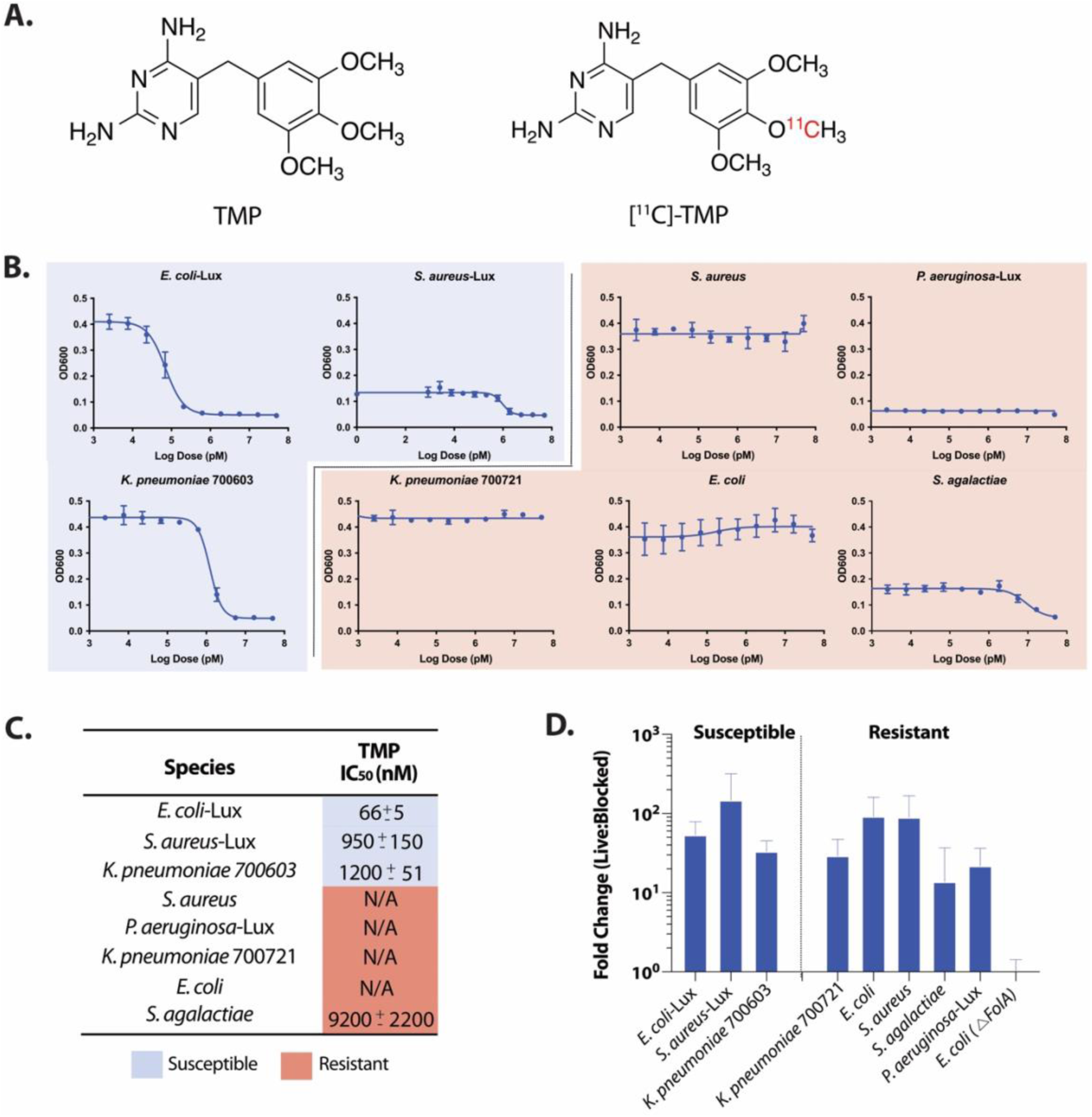
Structure of [^11^C]-TMP and *in vitro* TMP dose-response assays of different bacterial strains. **A)** Structures of Trimethoprim (TMP) and [^11^C]-TMP. **B)** TMP dose-response assay on bacterial strains. OD600 measurement was taken following a 6-hour incubation of different bacterial strains with TMP. **C)** Table summarizing IC_50_ values for TMP in different bacterial strains. The susceptibility or resistance of a bacterial strain to TMP is color-coded based on the IC_50_. **D)** Fold change of [^11^C]-TMP uptake relative to excess unlabeled TMP in bacterial cultures after a 30-minute incubation at 37°C (50 μM). Errors bars, SD (n=5).

[^11^C]-TMP uptake was tested in both TMP-susceptible and TMP-resistant bacteria by incubating the strains for 30 minutes with the radiotracer. Heat-killed or excess unlabeled TMP-blocked conditions served as controls. A DHFR knockout strain of *E. coli* K549 (ΔfolA) was tested as an additional control. All uptake values were normalized to the uptake values of the corresponding strain’s blocked control for comparison across species. High uptake was seen across all strains, except for the DHFR knockout *E. coli* K549 (ΔfolA), regardless of their susceptibility to TMP (**Fig. 1D**). TMP-resistant *K. pneumoniae* 700721, *E. coli, S. aureus, S. agalactiae*, and *P. aeruginosa* showed uptake 33-fold, 106-fold, 103-fold, 16-fold, and 25-fold higher than DHFR knockout *E. coli*, respectively. Similar results were observed when comparing uptake to heat-killed controls (**Fig. S2**). The retained [^11^C]-TMP uptake in all strains was promising and suggested that trimethoprim resistance was not simply the mutation of the TMP binding site of the endogenous DHFR gene, and that more investigation was warranted.

Resistance to TMP is often mediated by altered gene regulation, leading to increased translation of the DHFR enzyme as well as horizontal gene transfer of mutant DHFRs to which TMP binds poorly (*13*). To elucidate the resistance mechanisms present in our panel of bacteria, we performed whole-genome sequencing (WGS) on the following TMP-resistant subset of strains: *E. coli, K. pneumoniae* 700721, *S. agalactiae, P. aeruginosa*, and *S. aureus*. The genomes were assembled using an A-Bruijn assembler, and the completeness of the genomes was assessed using sets of single-copy genes within a phylogenetic lineage (*14, 15*).

Following quality control, predicted DHFR open reading frames (ORFs) from the assembled genomes of all five strains were individually investigated using the NCBI’s BLASTp database and literature search (**Fig. 2A**). *S. aureus* and *E. coli* each contained two different DHFR genes. In both cases, one gene was a resistant DHFR enzyme termed *DfrA*, which mediates TMP resistance (*16, 17*). The second gene was the canonical, wild-type (WT) DHFR for the strain that is known to bind TMP. *K. pneumoniae* similarly carried a WT DHFR known to bind TMP (*18*) as well as a DHFR gene whose TMP binding, to the best of our knowledge, is uncharacterized to date. However, only one ORF was found in *P. aeruginosa*. UniProt and NCBI databases suggest this is the native DHFR gene. It has been shown that drug-resistant strains of *P. aeruginosa* express export pumps that confer multi-drug resistance to TMP and other antibiotics (*19, 20*), which was apparent in our dose response assays (**Fig. S1**). Of note, previous reports showed when drug-susceptible and resistant strains of *P. aeruginosa* were lysed, TMP inhibited DHFR enzyme catalytic activity of all strains with equal potency (*21*), suggesting that P. aeruginosa DHFR may still bind to TMP radiotracers. For *S. agalactiae*, only one DHFR was found in the genome, with no reports on the whether or not it confers resistance.

**Figure 2.**
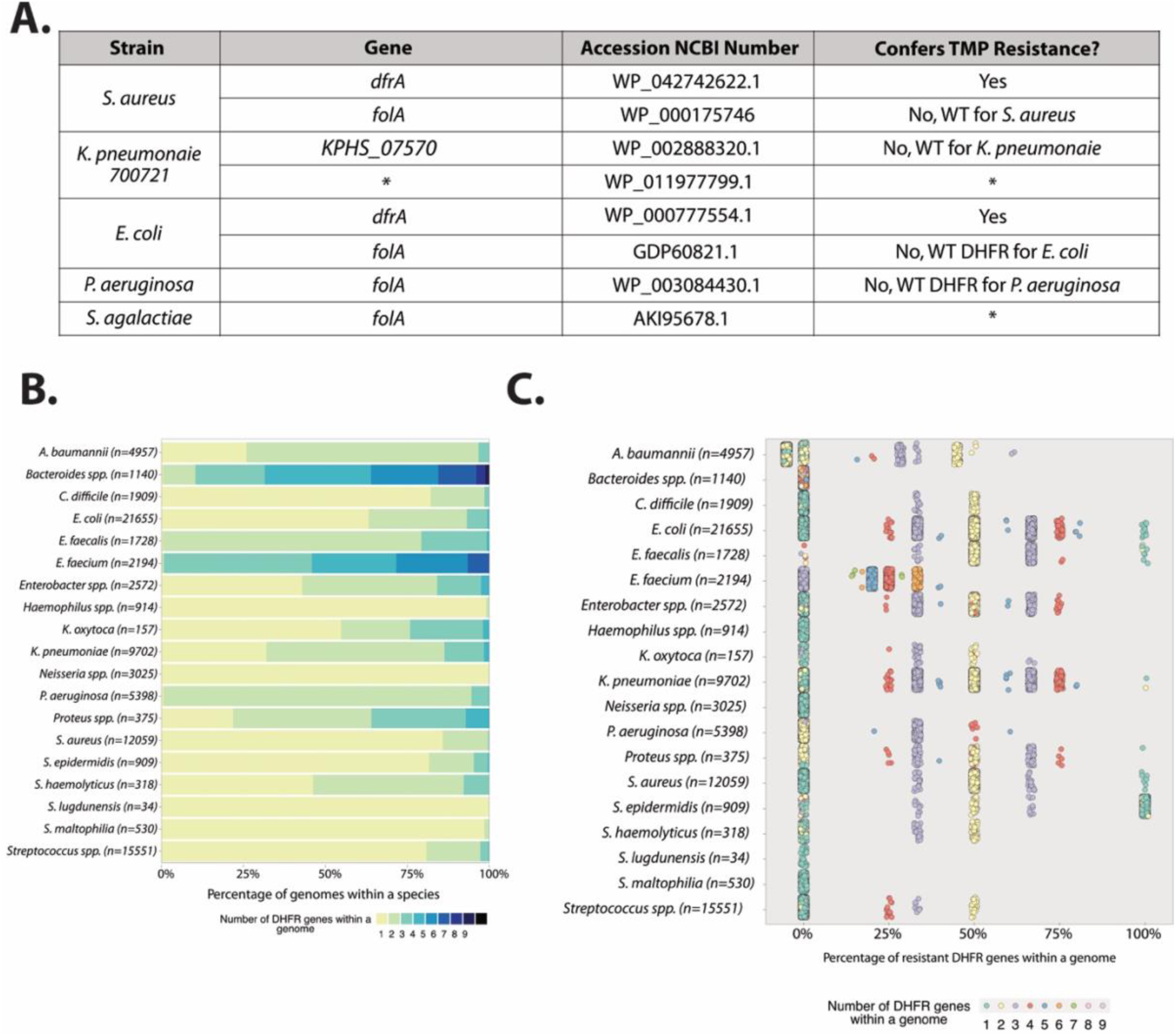
Whole genome sequencing characterization of lab and clinical isolates strains and bioinformatic analysis of clinically relevant bacteria. **A)** Table of DHFR open reading frames showing that most strains carried a DHFR copy known to bind TMP.* Denotes no literature found on these DHFR genes **B)** Proportion of clinically relevant bacterial strains from the NCBI RefSeq database with the indicated number of DHFR genes per genome. **C)** Resistance characterization of DHFR genes in relevant bacterial strains from NCBI RefSeq database. Each dot represents a strain.

It has been shown that the DHFR redundancy can be a common feature of clinically relevant bacteria (*22*). Given the potential implications for clinical imaging with TMP radiotracers and our observations from WGS of several strains which maintained TMP uptake despite resistance, we broadly surveyed the DHFR genes in the NCBI RefSeq deposited genomes of the top 19 most clinically relevant bacterial pathogens (**Fig. 2B**). We found that there is high heterogeneity both between and within species regarding the number of DHFRs carried. For example, the five most common bacterial pathogens responsible for healthcare-associated infections are *E. coli, S. aureus, K. pneumoniae, P. aeruginosa*, and *E. faecalis (23*). These strains on average carry 1.44, 1.14, 1.84, 2.05, and 2.21 DHFRs per genome, respectively. The percentage of resistance-conferring DHFR genes was calculated using the gene annotation. We found that almost always there is a native DHFR within these pathogenic species. In fact, only 0.56% of strains exclusively carried DHFRs that confer TMP resistance, a total of 742 out of 132,878 strains assessed. Moreover, we show a pattern relating the number of total DHFR genes per genome and the proportion of those genes that are TMP resistant (**Fig. 2C and S6-7**). Taking *E. coli* as an example, when a strain bears only one DHFR (cyan dots), that single gene is not resistant in almost all cases. Conversely, when a strain bears two DHFRs (yellow dots), typically one out of the two is resistant (50%). Furthermore, when strains bear three DHFRs (purple dots), typically either one and two DHFRs (33% and 67%), confer resistance. The pattern holds for four and five DHFRs, and so on (**Fig. S6**). Thus, it is reasonable to assume that [^11^C]-TMP has the potential to image TMP-resistant infections in different clinical settings, with a potential exception of the rare bacterial strain that has a lone copy of a TMP resistant DHFR gene.

Based on our pre-clinical evaluation of TMP radiotracers in animal models of infection and these promising data, we developed a clinical protocol to assess the biodistribution of [^11^C]-TMP in human subjects. Patients were consented for investigational protocol (NCT03424525) and several case examples are presented here. Firstly, a biodistribution study in a male in his 60’s that was being surveilled with [^18^F]-FDG PET/CT for metastatic lung adenocarcinoma was performed (**Fig. 3**, left panel). [^18^F]-FDG, a fluorinated glucose derivative, is often used for cancer patients to stage or monitor therapy as tumors are highly metabolically active. The diagnostic challenge with [^18^F]-FDG is that both cancer and infection often show elevated levels of FDG uptake. The patient had several sites of [^18^F]-FDG avid lung metastases that maintained low-level background uptake on the [^11^C]-TMP scan, suggesting that tumor metabolism does not confound [^11^C]-TMP uptake. Conversely, in a patient with known chronic lung infection, a woman in her 40’s with cystic fibrosis (CF), there were multiple foci of radiotracer uptake in the lungs corresponding with areas of multifocal pneumonia (**Fig. 3**, right panel). Notable sites of [^11^C]-TMP metabolism and excretion in patients included the liver, kidneys, and bladder, while many tissues that could be potential sites of infection showed very low background uptake. Radiotracer uptake was noted in vertebral bodies and proximal long bones for patients with metabolically active marrow (e.g. young women), and the time activity curves of select vertebral bodies were calculated for several patients showing that the [11C]-TMP uptake in bones does not increase over time (**Fig. S9**).

**Figure 3.**
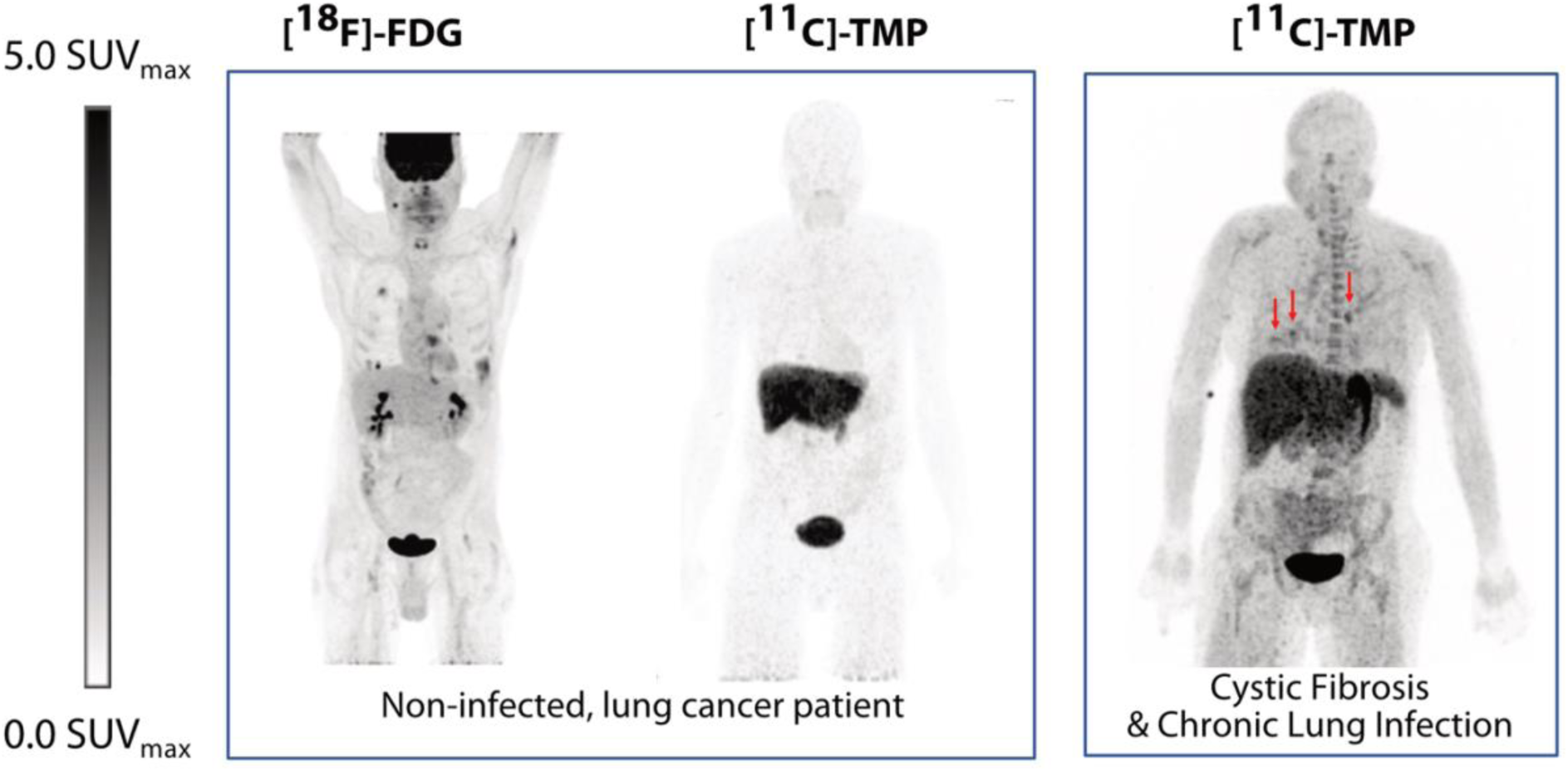
Biodistribution of [^18^F]-FDG vs [^11^C]-TMP in a patient with lung cancer and biodistribution in a patient with underlying chronic lung disease. The left-panel shows a man in his 60’s with known lung adenocarcinoma who underwent a [^18^F]-FDG (549 MBq) then a [^11^C]-TMP (563 MBq) PET/CT two days later. Whole body maximum intensity projection (MIP) images demonstrate the difference in biodistribution of the tracers. In the lungs, [^18^F]-FDG is taken up both by metabolically active tumor and inflammatory cells, whereas [^11^C]-TMP is not. Right-panel shows a comparison MIP image of a patient with cystic fibrosis and chronic lung infections who underwent a [^11^C]-TMP PET/CT (487 MBq). The right-panel patient is a woman in her 40’s with several foci of infection in the chest (red arrows). Other sites of signal include the liver, the kidneys, red bone marrow, and the stomach.

Further analyses of the female patient in her 40’s with CF shown in the right panel of Figure 3 determined that the lesions in the lungs had varying time-activity curves. Several lesions showed increased uptake over time (**Fig. 4A**). This uptake contrasted with the washout kinetics of the muscle, lymph nodes, and aorta (blood pool) measurements. Sputum cultures around the time of initial imaging from this patient grew cephalosporin-susceptible, but TMP-resistant *E. coli* (in addition to pan-resistant *Achromobacter*). She was placed on ceftriaxone IV for 2 weeks, and a follow up [^11^C]-TMP scan at the end of treatment (4 weeks after the first PET/CT) showed improvement in the right lower lobe foci on both PET and CT imaging. However, a new focus developed in the para-aortic, medial left lung, SUV_max_ 4.0 (**Fig. 4B**). A repeat sputum culture at the time of follow up imaging grew methicillin- and TMP-sensitive *S. aureus* in addition to her chronic *Achromobacter*. Thus, the radiotracer continued to show increased uptake in the setting of positive sputum cultures and despite antibiotic treatment.

**Figure 4.**
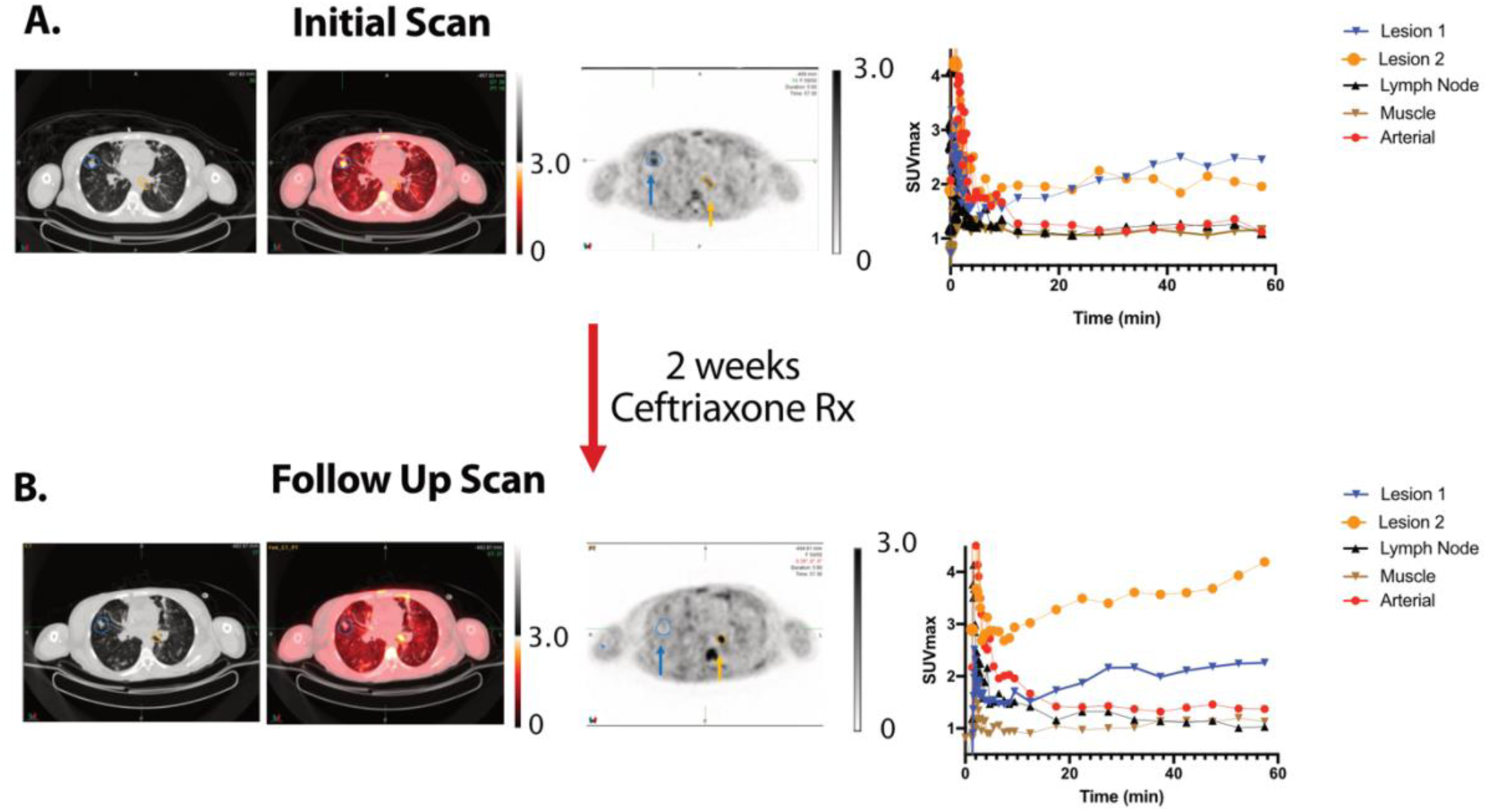
Time activity curves and bacterial heterogeneity. [^11^C]-TMP PET/CT images were acquired of a woman in her 40’s with an acute exacerbation of cystic fibrosis before **A)** and after treatment with 2 weeks of intravenous ceftriaxone **B)** Regions of interest were drawn around two separate pulmonary airspace opacities. In addition, reference regions were also drawn around a reference lymph node, paraspinal musculature, and within the aorta. Comparing the PET/CT images before and after treatment, the visible changes in relative [^11^C]-TMP uptake in lesion 1 versus lesion 2 demonstrate the bacterial heterogeneity and an apparent new infection with *S. aureus* based on sputum cultures. Patient received 487 MBq and 780 MBq of [^11^C]-TMP at the first and second time points, respectively.

In a companion case, a female patient in her 20’s also with CF was scanned 2 days into an inpatient course of intravenous antibiotics. Her scan showed an area of left lung consolidation with increased [^11^C]-TMP uptake (**Fig. 5**). This patient’s lung function and symptoms improved on antibiotics, and she was discharged from the hospital without a follow up [^11^C]-TMP PET/CT scan.

**Figure 5.**
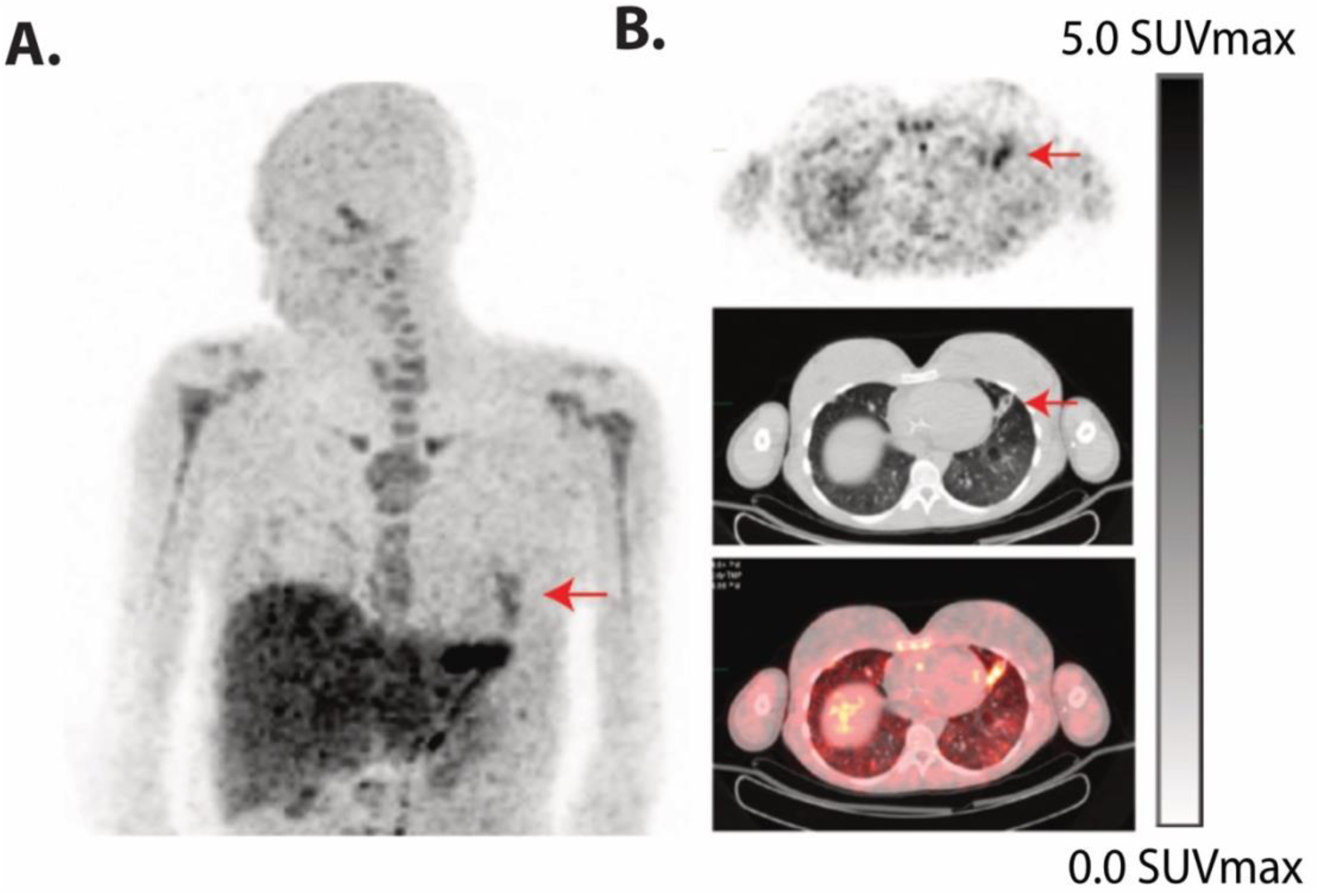
Acute exacerbation of cystic fibrosis. **A)** [^11^C]-TMP PET/CT (325 MBq) images were acquired of a female patient in her 20’s with cystic fibrosis and a background of known multifocal opacities, who presented with symptoms of an acute exacerbation. Patient was on intravenous antibiotics at the time of imaging. **B)** Coronal MIP and axial PET/CT images show a distinct lingular lesion with abnormal [^11^C]-TMP uptake, indicating the site of superimposed pneumonia.

Finally, a male patient in his 50’s with biopsy proven methicillin-sensitive *S. aureus* (MSSA) discitis osteomyelitis with contiguous involvement of the left L4-5 facet and surrounding musculature was scanned with [^11^C]-TMP. The patient underwent different scanning protocols (biodistribution versus kinetic) at the two treatment time points, having differences in radiotracer dosage and acquisition timing. To optimally compare the two studies, the treatment follow up dynamic scan images were reconstructed at 8 and 27 minutes post-injection to match the initial biodistribution imaging time points (**Fig. 6A**). After 6 weeks of intravenous cefazolin therapy, there was resolution of radiotracer uptake at the left L4-5 facet. These functional imaging findings preceded the anatomic sequelae of infection and remodeling seen on CT during the treatment and post-treatment interval (**Fig. 6B**). Notably, despite resolution of radiotracer uptake and no subsequent clinical recrudescence of infection, an MRI scan post-therapy suggested interval worsening of discitis/osteomyelitis (**Fig. 6C**). This discordance highlights the potential for [^11^C]-TMP to monitor active infections and provide a more accurate imaging surrogate for the resolution of infections, where standard imaging modalities may be ambiguous.

**Figure 6.**
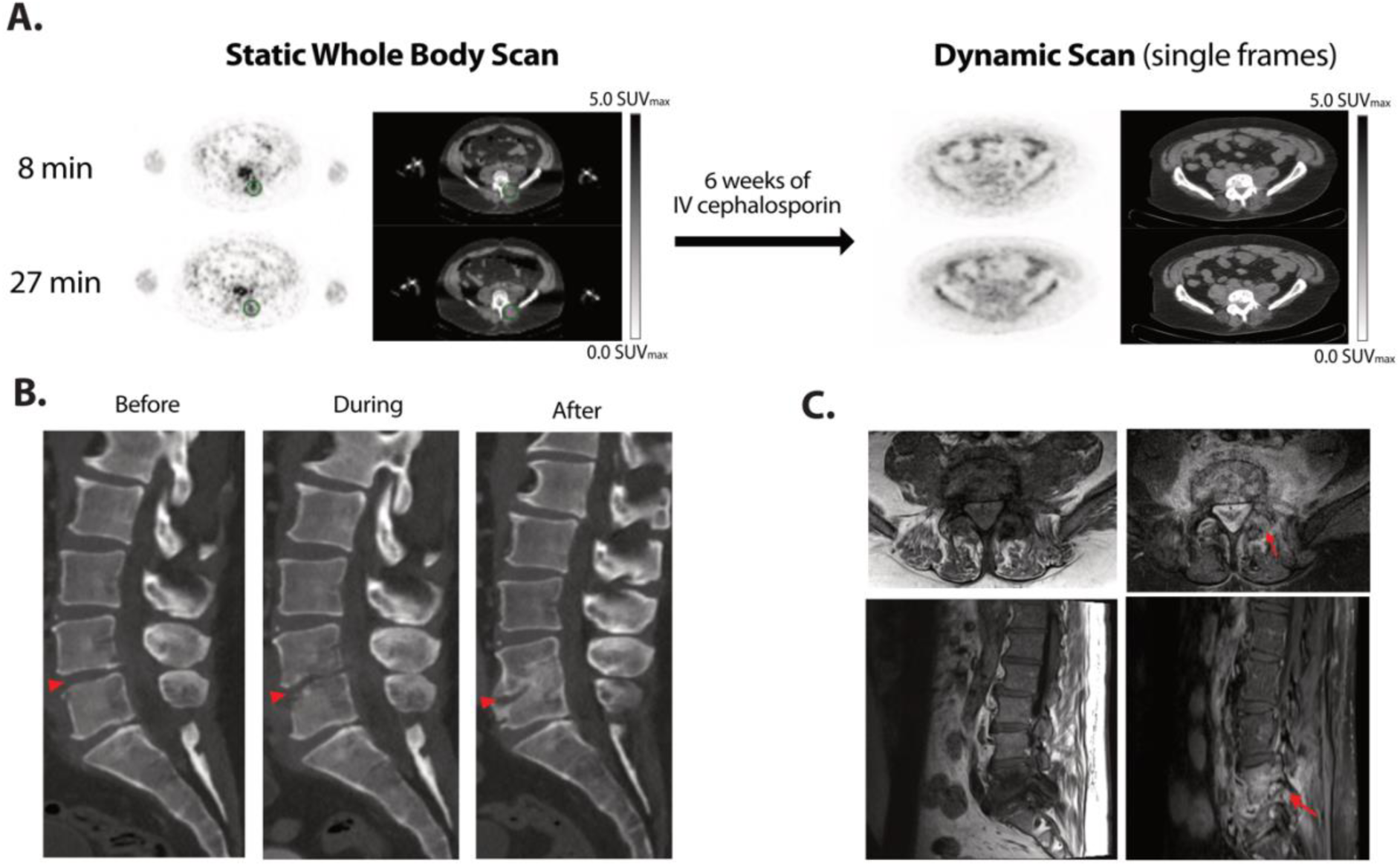
Biopsy proven discitis-osteomyelitis treated with antibiotics. A man in his 50’s with clinically suspected lumbar discitis-osteomyelitis was scanned with [^11^C]-TMP PET/CT at the initiation of empiric antibiotic therapy and after 6 more weeks of targeted antibiotic therapy. **A)** Axial PET/CT images show a clear site of asymmetric [^11^C]-TMP uptake in the left L4-5 facet at the start of therapy and lack of uptake after 6 weeks of intravenous treatment. Facet biopsy of the left L4-5 facet grew *methicillin-sensitive S. aureus*. Note, the patient received different doses of [^11^C]-TMP, 129 MBq at the first time point and 672 MBq at the second time point, thus the image quality was noisier at the first time point. **B)** The temporal morphologic sequelae of discitis-osteomyelitis are demonstrated by sagittal CT images before, during, and after treatment. **C)** In contrast to the PET/CT images, the gadolinium-enhanced MRI images of the patient at 10 weeks post therapy continue to demonstrate marrow replacement and contrast-enhancement, findings that are nonspecific for active infection versus continued inflammation.

## Discussion

Although bacterial infections often are treated effectively by antibiotic therapy, the incidence of multidrug resistant strains continues to rise and have a profound impact on modern medical care. Our diagnostic armamentarium for bacterial infections needs to be improved and molecular imaging can greatly contribute to the effort. Nuclear imaging, especially with anatomic correlation via CT or MRI, has the sensitivity to detect infections in human subjects. Here, we describe the assessment of a TMP radiotracer in antibiotic susceptible and resistant bacterial strains, catalog many of the most pathogenic strains of bacterial with respect to their potential imageability, and provide several case examples of first-in-human [^11^C]-TMP imaging.

We sought to understand whether antibiotic resistance would abrogate antibiotic tracer uptake. We found that drug-resistant bacterial species had similar uptake of [^11^C]-TMP as non-resistant species (**Fig. 1D**), suggesting that antibiotic resistance alone was not a critical feature impacting radiotracer uptake. Given that [^11^C]-TMP uptake was maintained in the resistant strains, we performed WGS and identified that three resistant strains carried a second DHFR gene, with the WT copy maintained. Taken together, these results suggested that the maintenance of at least one WT copy of DHFR was a critical step needed to maintain imageability.

Next, we cataloged the annotated DHFR genes from 19 bacterial species that are common causes of pathologic human infections, leveraging decades of research in antibiotic resistance mechanisms and genomic data (*24, 25*). We found that RefSeq deposited strains carried anywhere from 1-9 copies of DHFR (**Fig. 2B**). Outliers among the species included *E. faecalis* and *P. aeruginosa* where almost all strains analyzed contained multiple copies of DHFR, whereas *S. ludgunensis, Haemophilus spp*. and *S. maltophilia* strains mostly contained a single copy of DHFR. Bacteria such as *Bacteroides spp* and *E. faecium* had the most redundant copies of DHFR.

Almost all strains contained at least 1 copy of WT DHFR. For example, only 742 strains out of 132,878 strains assessed contained resistant DHFR copies only. That equals 0.56% of potential pathogenic bacteria included in our search, suggesting a strong potential for [^11^C]-TMP to detect many different bacterial strains in patients, assuming other criteria such as bacterial density and background tissue uptake are not limiting.

Interestingly, we see strong *in vitro* uptake in bacteria expressing drug export pumps. Several of these bacteria (*S. aureus, K. pneumoniae, E. coli, P*.*aeruginosa*) that underwent WGS showed drug export pumps (**Fig. S10**), yet, even *P. aeruginosa* showed [^11^C]-TMP uptake to be maintained. We hypothesize that the intracellular concentration of tracer needed to image a strain may be significantly less than the amount of drug needed to exert antimicrobial effects. Thus, these *in vitro* assays and genomic data set suggest that antimicrobial tracer binding is not categorically related to strain antibiotic resistance.

To understand the background uptake in terms of biodistribution and present early experience with [^11^C]-TMP in patients with suspected bacterial infection, we developed a first-in-human clinical protocol. The biodistribution of [^11^C]-TMP is different than that of the commonly used metabolic radiotracer [^18^F]-FDG (**Fig. 3**). Due to the remarkably low background radiotracer uptake in many tissues (i.e. the lungs, muscles, brain, and vasculature), the sensitivity of [^11^C]-TMP to detect acute bacterial infection is promising. Organs with the most radiotracer uptake in non-infected patients were the liver, kidneys, and bladder, which are the expected organs of metabolism and excretion. Future studies including [^11^C]-TMP dosimetry and kinetic modeling are nearing completion.

The lung has a naturally low background of [^11^C]-TMP. At the same time, patients with CF should have a high chronic bacterial burden in the lungs. We found that patients with lung infections had increased uptake in some but not all of their lung lesions as identified on CT imaging (**Figs. 3-5**). Lesions that show greater uptake may be more likely to be the cause of the patient’s active symptoms. This uptake could be monitored over time and relative to antimicrobial therapies. Alternatively, we are considering whether [^11^C]-TMP has the potential to be an additional biomarker that could be used in concert with clinical symptoms, biochemical lab values, and pulmonary function tests to stratify patients that may be candidates for lung transplantation. For example, a patient with advanced CF showed the development of a new focus of [^11^C]-TMP uptake while also demonstrating a new MSSA bacterial lung infection on sputum culture (**Fig. 4**). One lesion showed increased uptake on a time-activity curve, suggesting continued localization of the tracer to the infection (**Fig. 4B**).

Finally, we present a case of L4-5 vertebral discitis osteomyelitis with a biopsy and bone culture that grew MSSA. The uptake that was seen associated with the facet and nearby soft tissue resolved after 6 weeks of IV antibiotic therapy, suggesting the potential of such bacterial imaging radiotracers to monitor infection. Interestingly, the bony changes on CT were poor surrogates for active infection, and the enhancement on MRI suggested continued osteomyelitis long after the patient had completed antibiotics and the [^11^C]-TMP uptake had resolved, which supports adding [^11^C]-TMP imaging to monitor infections in some select clinical scenarios.

A limitation of our study is in the number strains of bacteria tested *in vitro*. It is possible that some strains could have significantly less uptake than others and that low level of uptake would portend those strains to be more difficult to detect *in vivo*. Further studies in animals, especially in non-human primates, may be helpful to best characterize such thresholds as rodents have different immune systems, metabolic rates, and imaging constraints. Another caveat that is evident for [^11^C]-TMP imaging is that there is increased uptake at the early time points in metabolically active marrow, seen best in vertebral bodies and proximal long bones in young patients (**Fig. 5**), however uptake does not increase over time (**Fig. S9**). This finding is not surprising as sustained therapeutic dose TMP is known to suppress hematopoietic activity. We suspect that this may limit the use [^11^C]-TMP at early time points, and given the physical half-life of ^11^C dominates washout period (biologic half-life), longer lived TMP radiotracers such as [^18^F]-fluoropropyl-trimethoprim are in development (*10*).

In summary, we present the testing of bacterial uptake of [^11^C]-TMP in bacteria that are both sensitive and resistant to TMP, describe the mechanism of TMP resistance in clinical isolates using WGS and apply a bioinformatic approach to highlight the potential for TMP to image different pathogens regardless of resistance status. We also demonstrate several case examples of patients with proven infections. Future studies describing the imaging methodology, kinetics, dosimetry, and additional patient examples are in progress and the studies presented here lay the foundation for future work characterizing the sensitivity and specificity of the TMP radiotracer family for bacterial indications.

## Materials and Methods

| Strain | Source |
| --- | --- |
| *E. coli*-Lux<br>(*E. coli* (GR12) EM7-lux::Km<br>[kanamycin resistant]) | Gift from Chris Contag Laboratory, Stanford University, Stanford, CA |
| *S. aureus* Lux<br>(*S. aureus* (8325-4; P2) lux::Cm<br>[chloramphenicol resistant]) | Gift from Chris Contag Laboratory, Stanford University, Stanford, CA |
| TMP/SMX Resistant *E. coli*<br>(Clinical Isolate) | Collaborator Laurel Glaser MD PhD,<br>University of Pennsylvania, Philadelphia, CA |
| TMP/SMX Resistant *S. aureus*<br>(Clinical Isolate) | Collaborator Laurel Glaser MD PhD,<br>University of Pennsylvania, Philadelphia, CA |
| *Klebsiella pneumoniae* subsp. *pneumoniae*<br>(Schroeter) Trevisan (ATCC® 700721™) | ATCC |
| *Klebsiella pneumoniae* subsp. *pneumoniae*<br>(Schroeter) Trevisan (ATCC® 700603™) | ATCC |
| TMP/SMX Resistant *S. agalactiae* (Group B<br>*Streptococcus*)<br>(Clinical Isolate) | Collaborator Laurel Glaser MD PhD,<br>University of Pennsylvania, Philadelphia, CA |
| *P. aeruginosa*<br>(Xen 5 (American Type Culture Collection<br>19660) lux::Tc<br>[tetracycline resistant]) | Gift from Chris Contag Laboratory, Stanford University, Stanford, CA |
| *E. coli* K549 (ΔfolA) | Gift from Elizabeth Howell's Laboratory,<br>University of Tennessee at Knoxville,<br>Knoxville, TN |

### [^11^C]-TMP Synthesis

[^11^C]CO_2_ was produced by ^14^N(p,α)^11^C reaction using an IBA Cyclone 18 (Louvain-la-Neuve), [^11^C]CH_3_I was synthesized from this using a gas-phase module (GE Healthcare). [^11^C]CH_3_I was trapped in a mixture of TMP-OH (0.75 mg, 2.70 mmol) and 5 N NaOH aqueous solution (5.4 μL, 27 μmol) in DMF (500 μL) at room temperature. The reaction mixture was heated at 70°C for 5 minutes and diluted with high performance liquid chromatography (HPLC) mobile phase (1.0 mL, 12% EtOH in 0.01 M phosphate buffer, pH = 3.0). The solution was then injected onto a HPLC equipped with a semi-preparative column (Phenomenex Gemini 5 *μ*, C18 110Å, New Column 250 × 10 mm) and eluted with HPLC mobile phase as above at a flow rate of 3 mL/min. The desired fraction eluted at 10-12 min was collected and used for biologic evaluation without concentration. For specific activity determination, an aliquot of [^11^C]-TMP was injected onto an HPLC equipped with an analytical column (Agilent XDB-C18, 5 *μ*, 150 × 4.6 mm) and eluted with a 15% CH_3_CN:85% water with 0.1% TFA at a flow rate of 1 mL/min (t_R_ = 5.1–5.2 min). Specific activity determinations were carried out by comparing the UV peak area (wavelength: 230 nm) of the desired radioactive peak with those of different concentrations of TMP by HPLC. An aliquot of [^11^C]-TMP was co-injected with TMP into an HPLC system to confirm its identity.

### Bacterial Cell Culture

Individual colonies were picked on Luria-Bertani (LB) plates with appropriate selection antibiotics. For experiments, all bacterial cultures were inoculated in LB broth with appropriate antibiotic selection and were shaken at 300rpm, 37°C overnight. For bioluminescent strains of bacteria, bioluminescence is not known to affect pathogenicity or drug uptake.

### In Vitro Assays

#### Bacterial Growth Inhibition-Dose Response Curves

All bacteria strains were grown overnight to saturation. On the day of the experiment, antibiotics solutions (Trimethoprim, Doxycycline, Ampicillin, and Chloramphenicol) prepared in LB broth were serially diluted 3:1 on a 96-well plate (with no drug control). 10 μL of the overnight bacterial cultures were diluted in 20 mL of LB and 10 μL of this diluted bacterial culture was added to each well. The plates were then incubated for 6 hours at 37°C while shaking at 180 RPM before measuring the OD600. Viability curves were plotted and analyzed on GraphPad Prism to determine an IC_50_.

#### [^11^C]-TMP Uptake Assays

All bacteria strains were grown overnight to saturation. On the day of the uptake experiment, OD600 of the bacteria was measured and used to determine the number of colony forming units (CFU). The cultures were sedimented by centrifuging at 4000 rpm and were resuspended at a concentration of 5×10^9^ CFU/mL in LB. Three, 5mL aliquots of each strain were prepared in tubes labeled as: “Live”, “Blocked”, or “Heat Killed”. Heat Killed aliquots were then heated to 95°C for 30 minutes while “Live” and “Blocked” bacteria were placed on ice.

Cold, unlabeled trimethoprim (TMP) was added to the “Blocked” aliquots to a final concentration of 50 μM. A radiotracer dose of 5×10^6^ counts per minute (CPM) was added to all aliquots of bacterial strains and were incubated at 37°C for 30 minutes. Following incubation, the cultures were centrifuged at 4000 rpm and washed twice with ice cold PBS. After the second PBS wash, the bacteria were again sedimented but then resuspended in 1 mL of PBS and split into 5 technical replicates of 200 μL. Radiotracer uptake was measured on Gamma Counter (Perkin Elmer) with decay correction.

Once the radiotracer had decayed (10 half-lives), a Lowry assay (Thermo Scientific) was run on each strain of bacteria to determine protein concentration. This protein concentration was used to normalize the radiotracer uptake to milligrams (mg) of protein. All analysis was performed using GraphPad Prism 9.

### Whole Genome Sequencing (WGS) for Identification of TMP Resistance Mechanisms

Bacterial cultures of TMP resistant strains (*E. coli, S. aureus, K. pneumoniae* 700721, *P. aeruginosa*, and *S. agalactiae* (Group B *Streptococcus)*) were inoculated in 5 mL of LB and shaken at 300 RPM, 37°C overnight. To prepare for whole genome sequencing, 1 μL of the overnight bacterial cultures were diluted in 100 μL of PBS.

DNA from cultured samples were isolated with the Qiagen DNeasy PowerSoil kit and libraries were generated using the Nextera Flex Library Prep kit and sequenced on the Illumina HiSeq 2500 using 2×125 bp chemistry.Illumina sequencing reads were demultiplexed and quality filtered using the default settings of Trimmomatic (*26*) and adapters were trimmed from sequences with Cutadapt software (*27*). Low complexity sequences were masked using Komplexity (https://github.com/eclarke/komplexity) with a normalized complexity score of < 0.55. Reads that mapped to a human reference sequence (Genome Reference Consortium Human Build 38, GRCh38) were identified using bwa (*28*) and reads with > 60% of the read fraction mapping to GRCh38 or with a percent identity > 50% were removed. This produced a mean of 1.02 million host-filtered, quality-controlled reads per sample. SPAdes 3.14 (*14*) was used for de novo assembly of the host-filtered, quality-controlled short reads. The quality of the assembled genomes was assessed using CheckM v1.1.2 (*15*) and Anvi’o v6.2 (*29*) for completion and contamination. Open reading frames of the assembled genomes were identified using Prodigal v2.6.3 (*30*) and blasted against the Comprehensive Antibiotic Resistance Database (CARD) v1.1.7 (*31*) and a manually curated list of DHFR genes. Pileup analysis was performed by aligning reads onto the assembled and reference WT DHFR genes using Bowtie2 (*32*) and counting the frequency of variants using dnapy (https://github.com/sherrillmix/dnapy).

### Survey of DHFR Genes in RefSeq Deposited Genomes

The protein fasta files of bacterial strains were downloaded with ncbi-genome-download (https://github.com/kblin/ncbi-genome-download). Using the searchdesc function from the okfasta package (https://github.com/kylebittinger/okfasta), DHFR and TMP-related genes were identified and saved separately. Next, R was used to parse through these DHFR fasta files and to count susceptibility or resistance to TMP based on the annotation provided by RefSeq.

### Patient Recruitment & Imaging Studies

The purpose of the study was to evaluate the biodistribution and kinetics of [^11^C]-TMP in human subjects. Patients with suspected or confirmed bacterial infections were screened against the inclusion/exclusion criteria. No power/sample size tests were calculated and the investigators were not blinded while the data was collected or analyzed. Written informed consent was obtained from all participants. All images and patient data are deidentified.

[^11^C]-TMP was prepared according to good manufacturing practices and as described above at the Penn Cyclotron Facility. Tracer was administered intravenously at doses listed in figure legends and an [^11^C]-TMP PET/CT was acquired on a Phillips Ingenuity PET/CT scanner (Philips Healthcare, Andover, MA) (*33*). Images were analyzed using MIM software (Cleveland, OH).

## Data Availability

The data that support the findings of this study are available from the corresponding author, MAS, upon reasonable request.

## Acknowledgments

We thank all patients that participated in this study. Additionally, we would like to thank the PennCHOP Microbiome program, the Penn Cyclotron, and PET Center for their invaluable expertise and infrastructure, all of which made this research possible. Finally, we would also like to thank Daniel Pryma for his helpful discussions.

## Author Contributions

I.K.L, D.A.J., and M.A.S. conceived of the project and experimental design. I.K.L., D.A.J., and O.A. performed experiments, analyzed data, and interpreted the results. V.T. and K.B. designed and performed bioinformatic analysis and interpretation. D.D. and D.H. referred patients for the clinical studies. J.K.C., A.Y., T.D. and R.D. analyzed patient scans. J.D.N., A.R., J.M.E., H.L., and R.H.M. were responsible for chemical and radiotracer synthesis. L.G. and R.M.K. contributed reagents, analysis, and interpretation. I.K.L., D.A.J., M.A.S., and J.C. wrote the manuscript with input from all authors.

## Data and Materials Availability

All data associated with this study are present in the paper or the Supplementary Materials.

