## Supplemental Information for "Imaging Sensitive and Drug-Resistant Bacterial Infection with [^11^C]-TMP: *In Vitro* and First-in-Human Evaluation"

Iris K. Lee[1†], Daniel A. Jacome[1†], Joshua K. Cho[1], Vincent Tu[3], Anthony Young[1], Tiffany Dominguez[1], Justin D. Northrup[1], Jean M. Etersque[1,2], Hsiaoju Lee[1], Andrew Ruff [1], Ouniol Aklilu[1], Kyle Bittinger[3], Laurel Glaser,[4] Daniel Dorgan,[5] Denis Hadjiliadis,[5] Rahul M. Kohli,[2,6] Robert H. Mach[1], David A. Mankoff[1], Robert Doot[1], and Mark A. Sellmyer[1,2]*

[1]Department of Radiology, University of Pennsylvania, Philadelphia, PA 19104, USA;
[2]Department of Biochemistry and Biophysics, University of Pennsylvania;
[3]Department of Gastroenterology, Hepatology and Nutrition, Children's Hospital of Philadelphia;
[4]Department of Pathology and Laboratory Medicine, Hospital of the University of Pennsylvania;
[5]Department of Medicine, Division of Pulmonary and Critical Care Medicine, University of Pennsylvania
[6]Department of Medicine, Division of Infectious Disease, University of Pennsylvania

† Co-first authorship

* Correspondence should be addressed to:
M.A.S.
https://orcid.org/0000-0002-1407-1905

**One Sentence Summary:** The PET radiotracer [$^{11}$C]-trimethoprim shows high uptake in both TMP-sensitive and -resistant bacteria *in vitro,* the potential for imaging many different pathogenic strains, and uptake in patients with active infections.

### Supplemental Figures

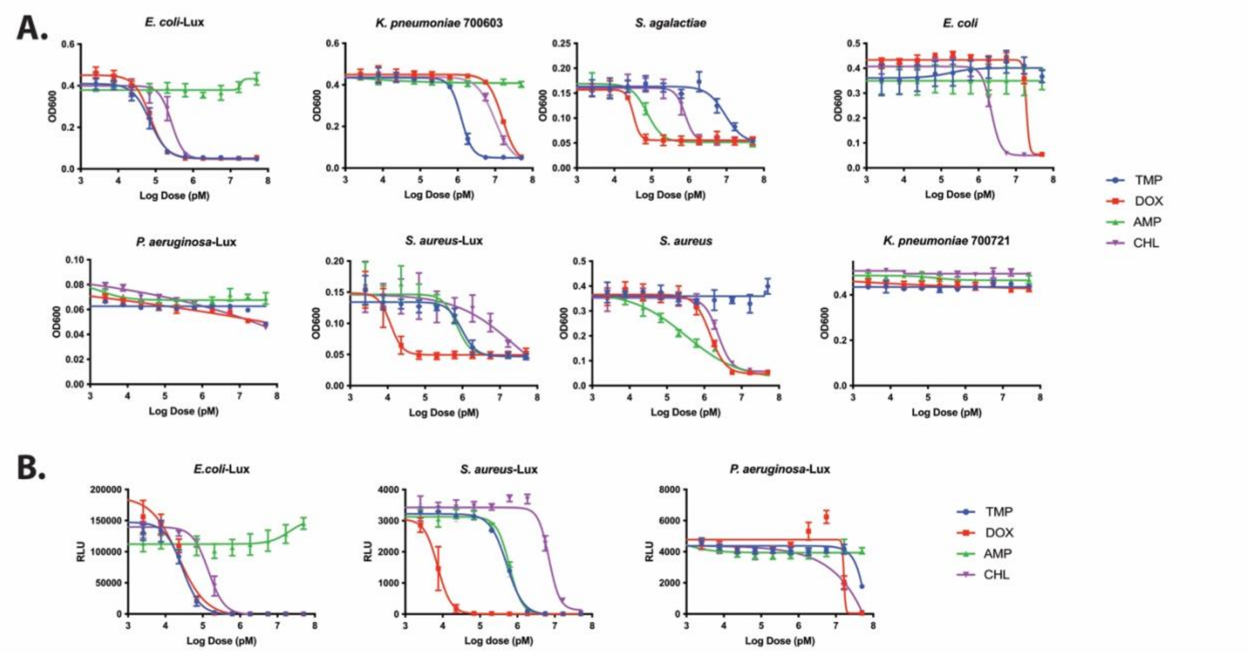


| Species | TMP | DOX | AMP | CHL |
|---|---|---|---|---|
| *S. aureus*-Lux | 950±150 | 12±2 | 700±100 | 23000* |
| *S. aureus* | N/A | 1430±120 | 320±28 | 2300±180 |
| *S. agalactiae* | 9100 ±2100 | 32 ±5 | 76±13 | 780 ±100 |
| *E. coli*-Lux | 66±5 | 68±5 | N/A | 250± 31 |
| *E. coli* | N/A | 19000* | N/A | 2200±120 |
| *K. pneumoniae 700603* | 1200± 51 | 16000* | N/A | 9800±670 |
| *K. pneumoniae* 700721 | N/A | N/A | N/A | N/A |
| *P. aeruginosa*-Lux | N/A | N/A | N/A | N/A |

Susceptible   Resistant

**Figure S1.** ***In vitro*** **characterization of susceptibility and resistance of different bacteria against a panel of common antibiotics. A)** Dose response assay of different bacterial strains with 4 different antibiotics. TMP = Trimethoprim, DOX = Doxycycline, AMP = Ampicillin, and CHL = Chloramphenicol. **B)** Luminescence readout of dose response assays from section A for bacterial strains expressing *lux*-operon. **C)** Table summarizing $IC_{50}$ of the antibiotics in different bacterial strains. *Denotes ambiguous curve on GraphPad Prism software

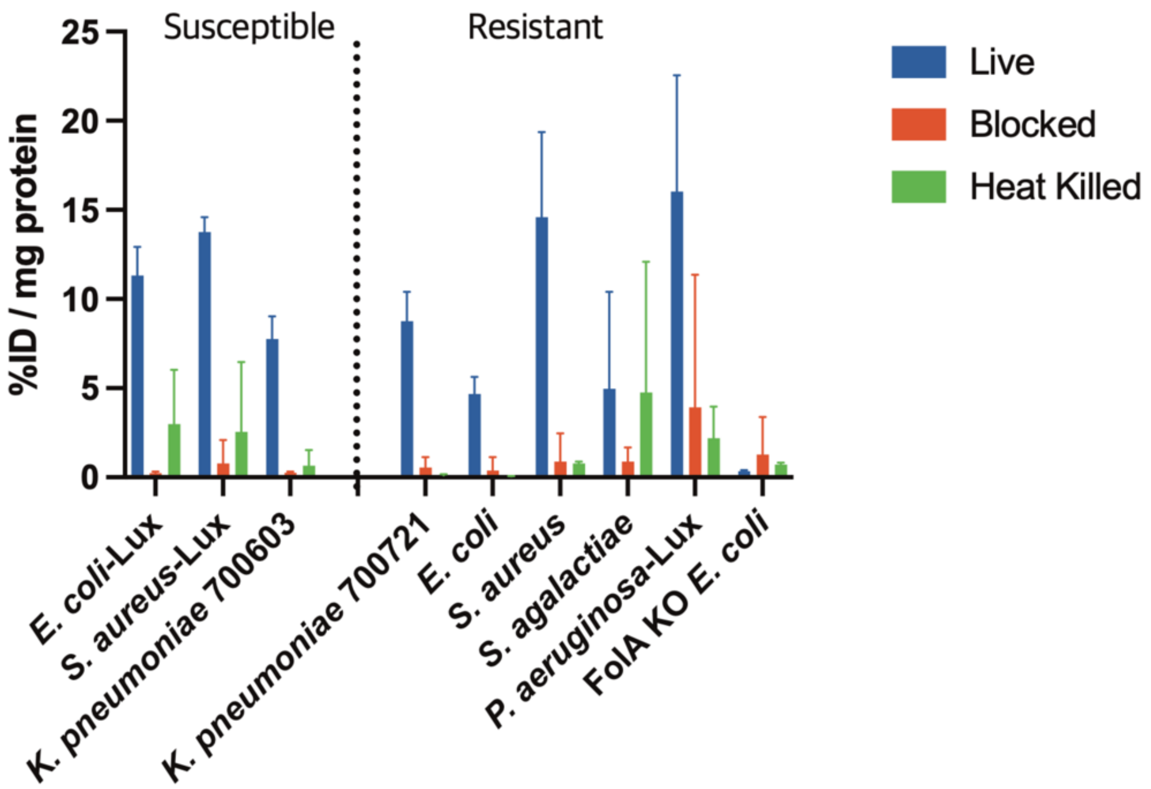


**Figure S2**. **Representative *in vitro* [$^{11}$C]-TMP uptake in bacterial strains**. [$^{11}$C]-TMP uptake in bacterial cultures after a 30-minute incubation with [$^{11}$C]-TMP. Controls were blocked with cold TMP (50 µM) or heat-killed at 95°C for 30 minutes. Mean percent injected dose per milligram protein (%ID/mg) plotted with errors bars in SD (n=5).

| Sample ID | Genome size (bp) | Contigs | Genes (#) | Completeness (%) | Contamination (%) | Strain Heterogeneity | $N_{50}$ (Scaffolds) | GC (%) | Mean Coverage |
|---|---|---|---|---|---|---|---|---|---|
| *E. coli* | 5097133 | 290 | 4917 | 99.56 | 0.43 | 0 | 119500 | 50.6 | 38.88 |
| *K. pneumoniae* 700721 | 5603052 | 459 | 5360 | 99.71 | 0.32 | 11.11 | 107184 | 57 | 29.95 |
| *P. aeruginosa* | 6666409 | 400 | 6446 | 99.54 | 0.55 | 11.11 | 45469 | 66.2 | 23.75 |
| *S. agalactiae* | 2073226 | 84 | 2095 | 99.78 | 0.13 | 0 | 63952 | 35.3 | 102.3 |
| *S. aureus* | 2781448 | 157 | 2617 | 99.11 | 0.32 | 0 | 50368 | 32.7 | 70.55 |

**Figure S3.** Completeness CheckM table

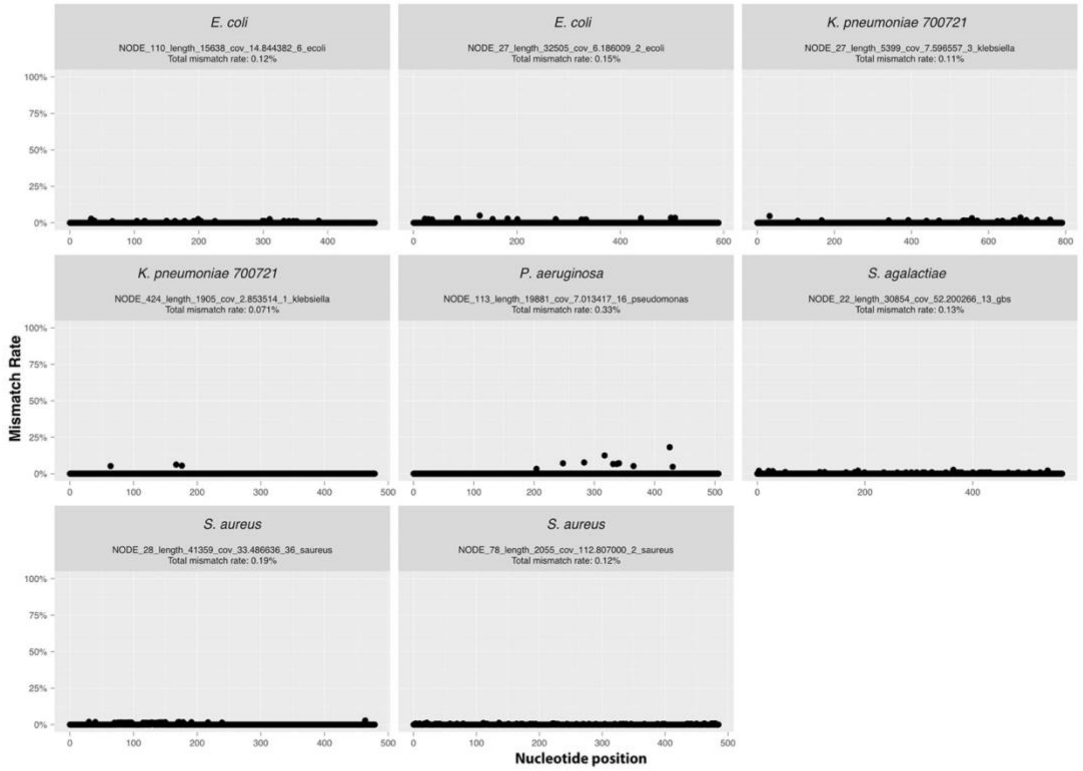


**Figure S4.** Pile up analysis on DHFR genes from assembled genomes.

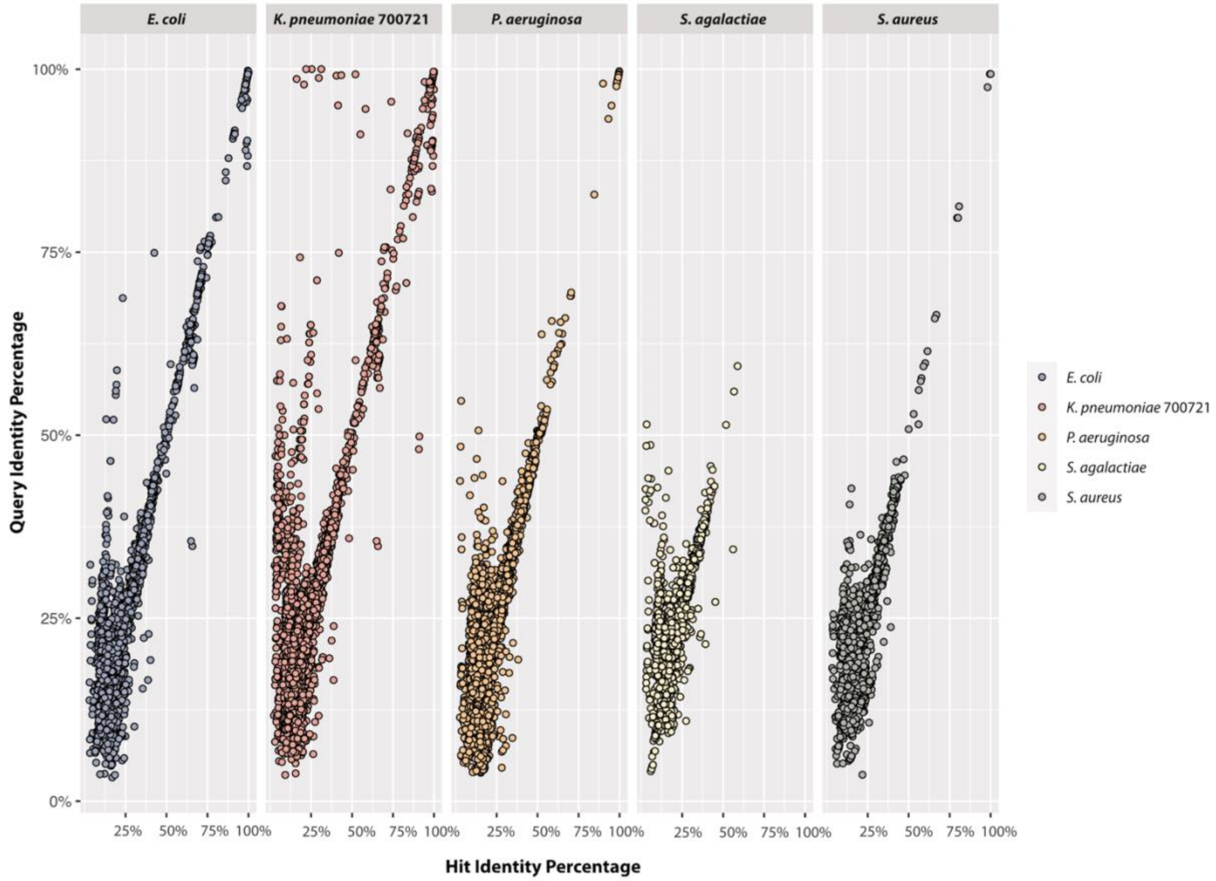


**Figure S5.** Blastp results relating assembled ORFs from the TMP-resistant subset of bacteria against different previously annotated antibiotic resistance genes in the Comprehensive Antibiotic Resistance Database.

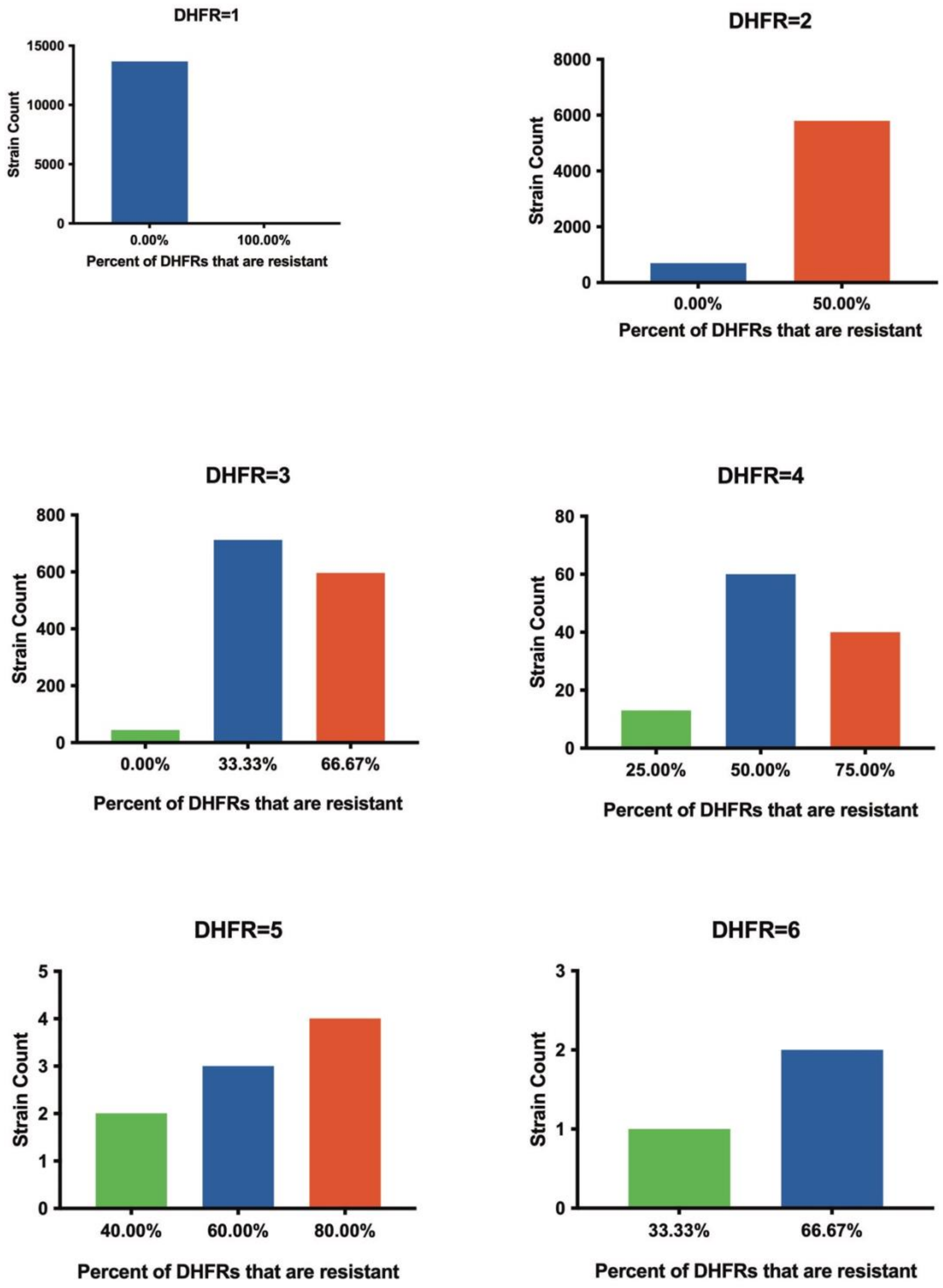


**Figure S6.** Distribution of resistance conferring DHFR genes based on total DHFR gene number within an *E. coli* strain.

**Percent of genes within a species that are resistant (%)**

**Number of DHFR genes within a strain**

| Species | 0% | 25% | 33.3% | 40% | 50% | 66.7% | 60% | 75% | 80% | 100% | Total |
|---|---|---|---|---|---|---|---|---|---|---|---|
| ***E. coli*** | **14416** | **13** | **713** | **2** | **5855** | **3** | **598** | **40** | **4** | **11** | **21655** |
| 1 | 13673 | | | | | | | | | 11 | 13684 |
| 2 | 699 | | | | 5795 | | | | | | 6494 |
| 3 | 44 | | 712 | | | | 596 | | | | 1352 |
| 4 | | 13 | | | 60 | | | 40 | | | 113 |
| 5 | | | | 2 | | 3 | | | 4 | | 9 |
| 6 | | | 1 | | | | 2 | | | | 3 |
| ***K. pneumoniae*** | **3597** | **17** | **614** | **4** | **4870** | **5** | **526** | **65** | **2** | **2** | **9702** |
| 1 | 3101 | | | | | | | | | 1 | 3102 |
| 2 | 465 | | | | 4804 | | | | | 1 | 5270 |
| 3 | 26 | | 614 | | | | 526 | | | | 1166 |
| 4 | 5 | 17 | | | 66 | | | 65 | | | 153 |
| 5 | | | | 4 | | 5 | | | 2 | | 11 |
| ***S. aureus*** | **10403** | | **18** | | **1610** | | **21** | | | **7** | **12059** |
| 1 | 10339 | | | | | | | | | 7 | 10346 |
| 2 | 59 | | | | 1610 | | | | | | 1669 |
| 3 | 5 | | 18 | | | | 21 | | | | 44 |

**Figure S7. Table showing expected pattern of resistance and DHFR genes within a single bacterial strain for select species.** The vertical axis displays the number of DHFR genes carried by the bacterial strain while the horizontal axis shows the percent of those genes carried that are resistant. The data in the central section of the table is the total number of genomes with the corresponding DHFR genes and resistance proportions.

| Inclusion Criteria |
| --- |
| **1.** At least 18 years of age |
| **2.** Known or suspected bacterial infection |
| **3.** Participants must be informed of the investigational nature of this study and provide written informed consent in accordance with institutional and federal guidelines prior to study-specific |
| **Exclusion Criteria** |
| **1.** Antibiotic therapy with trimethoprim within 48h of the baseline PET/CT scan |
| **2.** Inability to tolerate imaging procedures in the opinion of an investigator or treating physician |
| **3.** Serious or unstable medical or psychological comorbidities that, in the opinion of the investigator, would compromise the subject's safety or successful participation in the study |
| **4.** Females who are pregnant or breast feeding at the time of screening will not be eligible for this study; a urine pregnancy test will be performed in women of child-bearing potential at screening. |

**Figure S8.** Inclusion and Exclusion criteria for [$^{11}$C]-TMP trial.

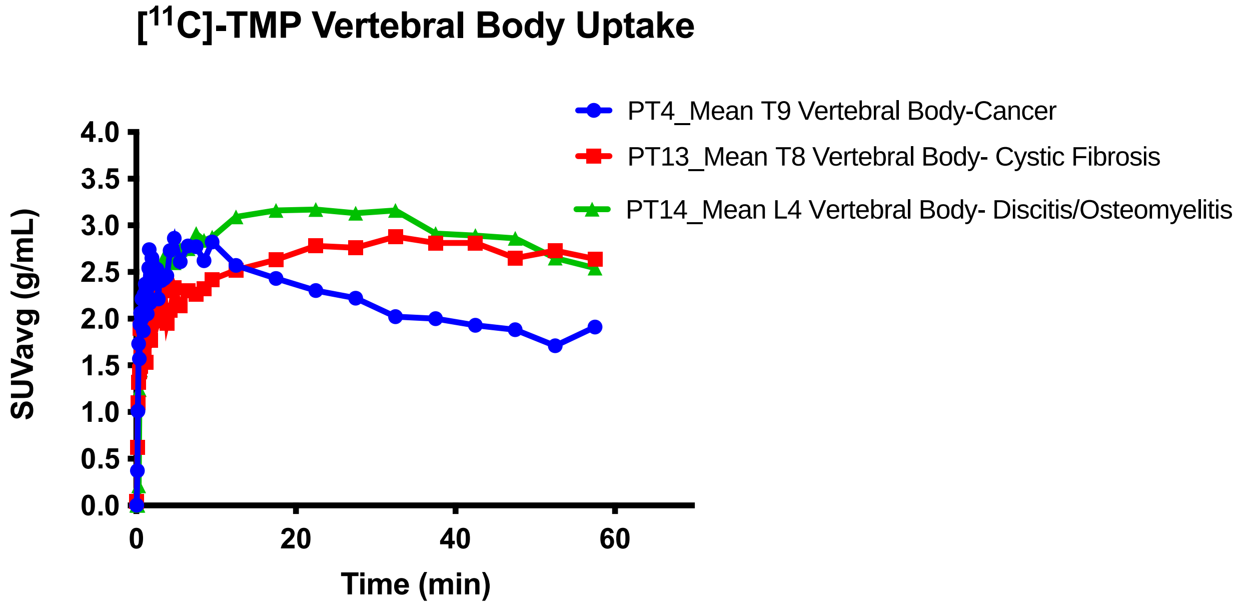


**Figure S9.** Vertebral Body Uptake for all patients as functions of time.

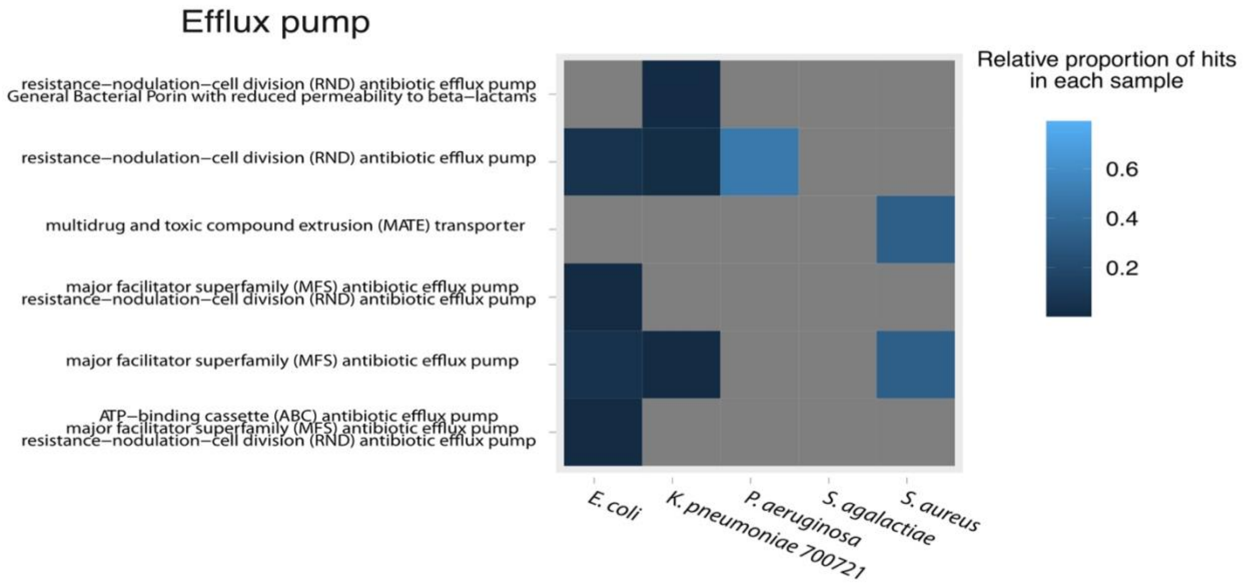


**Figure S10. Heatmap of efflux pump genes expressed in the panel of TMP-resistant bacteria.** Relative proportion is relative to the total number of genes within the species genome that had greater than 90% homology to Comprehensive Antibiotic Resistance Database annotated genes.
